# Estimation of the fraction of COVID-19 infected people in U.S. states and countries worldwide

**DOI:** 10.1101/2020.09.26.20202382

**Authors:** Jungsik Noh, Gaudenz Danuser

## Abstract

Since the beginning of the COVID-19 pandemic, daily counts of confirmed cases and deaths have been publicly reported in real-time to control the virus spread. However, substantial undocumented infections have obscured the true prevalence of the virus. A machine learning framework was developed to estimate time courses of actual new COVID-19 cases and current infections in 50 countries and 50 U.S. states from reported test results and deaths, as well as published epidemiological parameters. Severe under-reporting of cases was found to be universal. Our framework projects for countries like Belgium, Brazil, and the U.S. ∼10% of the population has been once infected. In the U.S. states like Louisiana, Georgia, and Florida, more than 4% of the population is estimated to be currently infected, as of September 3, 2020, while in New York the fraction is 0.12%. The estimation of the actual fraction of currently infected people is crucial for any definition of public health policies, which up to this point may have been misguided by the reliance on confirmed cases.

## Main Text

Since its initial spread in China in December 2019, the coronavirus disease 2019 (COVID-19) has caused more than 860,000 confirmed deaths all over the world as of September 3, 2020 (1), and it continues to threaten the whole population most of which remain susceptible to infection by the severe acute respiratory syndrome coronavirus-2 (SARS-CoV-2). As an effort to contain the virus, the daily counts of laboratory-confirmed cases and deaths have been publicly reported in real-time (2). However, substantial undocumented infections have obscured the actual fraction of at least once infected people. A computational study estimated the ratio of confirmed cases to actual cases, i.e., an ascertainment rate, to be only 14% during the early outbreak in China (3). Large-scale seroprevalence studies aimed to estimate the actual number of infections and found severe under-ascertainment in several U.S. states, where the ascertainment rates varied from 4.2% in Missouri to 8.9% in New York and 16.7% in Connecticut until March or April 2020 (4, 5). More importantly, we still do not know how many individuals are currently infected in many countries and regions. The currently infected population is the cause of future infections and deaths. Its actual size in a region is a crucial variable required when determining the severity of COVID-19 and building strategies against regional outbreaks.

This study presents machine learning-based estimates of actual sizes of currently infected populations in select countries and all 50 U.S. states. These fractions of infected people are derived by estimating daily ascertainment rates and subsequently adjusting the under-reported COVID-19 cases. The estimates are based on publicly available datasets of daily confirmed cases and deaths, and published estimates of key pandemic parameters. The dataset for countries is from the repository by the Center for Systems Science and Engineering (CSSE) at Johns Hopkins University (2), and the data for U.S. states are from the COVID Tracking Project (6). Since the developed pipeline requires simple input, it is widely applicable to more granular analyses of specific regions or communities, for which the number of confirmed cases and deaths are being tracked. Using the proposed pipeline, an online repository presents visualizations of daily updates on the estimated actual fraction of infected people for the 50 countries with the most confirmed cases and for all 50 U.S. states (7).

The daily counts of confirmed COVID-19 cases and deaths alone possess incomplete information on the relative abundance of epidemiological compartments of a population that is susceptible, infected, recovered, or deceased. Whether to be confirmed or not adds another layer of complexity to the categories of infected, recovered, or deceased compartments (Fig. 1). In addition to the under-ascertainment, several limitations in the reported data make it challenging to estimate the number of currently infected cases: recovery events are not tracked in most countries; there are time delays between infection onset and laboratory-confirmation (3); and even the death tolls are suggested to be under-reported in some regions (8, 9).

**Figure 1.**
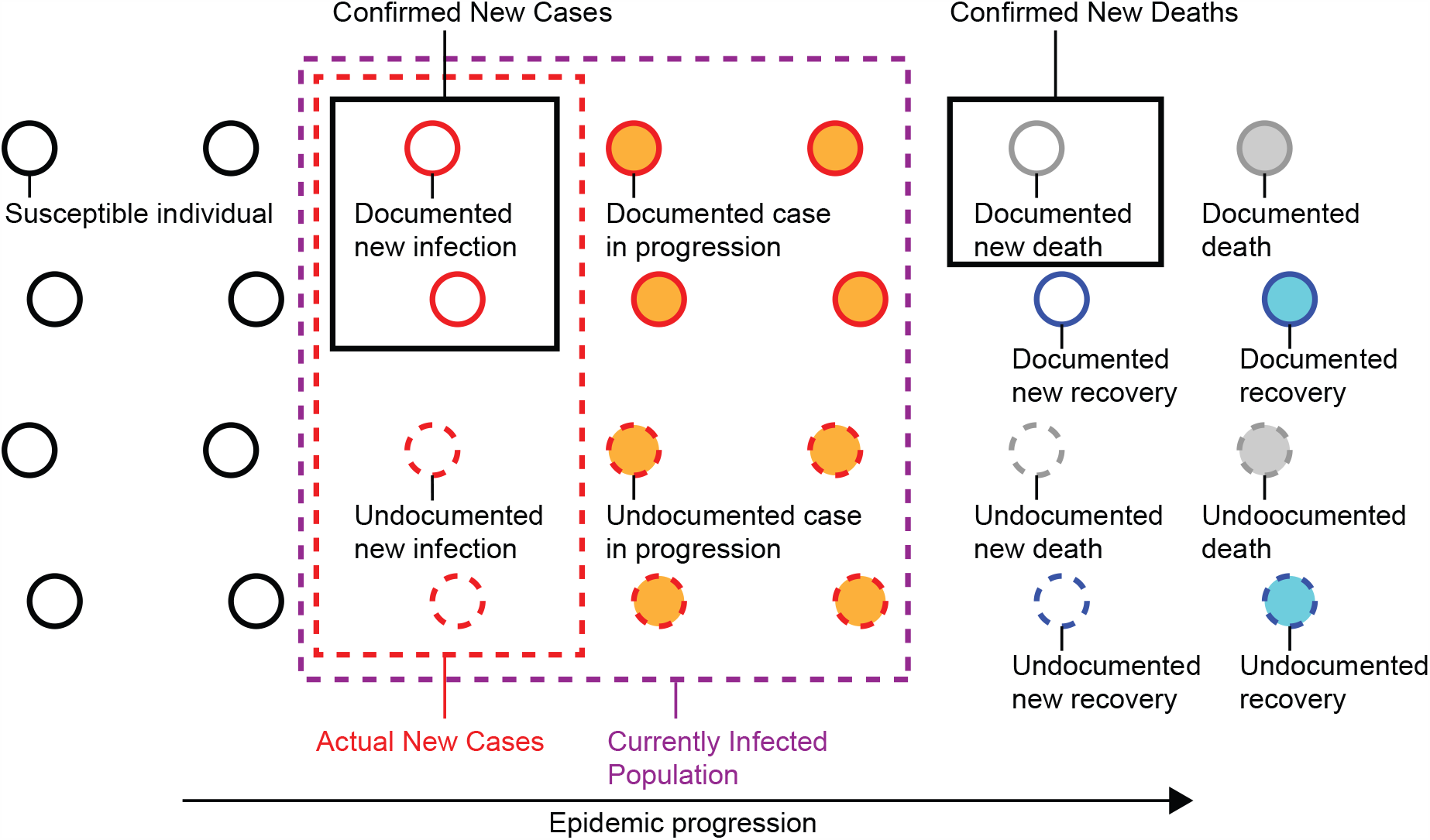
Undocumented COVID-19 cases. In an epidemic process, a population is categorized into susceptible, infected, deceased, or recovered individuals. Counts of confirmed COVID-19 cases, deaths, and recoveries are insufficient to calculate the number of currently infected individuals (purple dotted box) because of substantial undocumented infections not captured by diagnostic tests. The input to the proposed framework is the daily counts of confirmed new cases and deaths (black boxes). Using pandemic parameters such as the Infection-Fatality-Rate and the mean duration periods from infection to death and recovery, the framework estimates the counts of actual new cases (red dotted box) and currently infected individuals.

Key epidemiological parameters such as the Infection-Fatality-Rate (IFR) give us a clue to fill the gap between confirmed and actual infections, under the assumption that the number of undocumented deaths is negligible (Fig. 1). The IFR of COVID-19 has been a focus of intensive research, yet studies from different locations and times have not reached a consensus estimate (10). A recent large seroprevalence study in 133 cities of Brazil presented an IFR estimate of 1.0% (11). In a different approach, a study analyzed early pandemic data in China combined with the prevalence obtained by PCR-testing of the entire international resident population repatriated from China. The authors’ estimate of the IFR was 0.66% with a wide band of uncertainty (0.39%–1.33%, 95%-confidence interval) (12). The same study also reported the mean duration from onset of symptoms to death or recovery (17.8 and 24.7 days, respectively) based on individual-level data. These estimates were employed here to infer the actual number of infections across countries and regions. The IFR is known to heavily depend on age groups (12) and would vary across countries with different age distributions. Therefore, applying the above IFR estimate to a region with an extremely young or old population will be inappropriate. But, considering the estimate’s large estimation uncertainty, the confidence interval is expected to cover the true IFRs of most countries and U.S. states.

Our computational pipeline started with initial estimates of time courses of actual new infections and new recoveries, derived from the daily confirmed deaths, the IFR estimate, and the mean duration from infection to death and recovery (Fig. S1). The estimated new infections led to two other initial estimates: a daily ascertainment rate that is the ratio of confirmed new infections to the estimated new infections, and the number of currently infected cases each day. Then a regression model was applied to find a functional relation of the daily infected cases to the daily ascertainment rates, accounting for a common temporal trend of under-reporting shown in both of the daily ascertainment rate and the ratio of confirmed cases to infected cases. Employing the expectation-maximization (EM) algorithm, the pipeline iteratively updated the time courses of ascertainment rates, new infections, and currently infected cases based on each other until convergence to obtain final estimates. The same EM iterations were applied with the lower/upper limits of the IFR estimate to obtain upper/lower 95%-confidence limits of the estimated number of infections, respectively (Fig. S1, See Supplementary Methods for details).

The estimates of the actual number of infections were validated using seroprevalence data from the large-scale surveys conducted by the Centers for Disease Control and Prevention (CDC) and New York state (4, 5, 13). The surveys collected serum samples in multiple U.S. states and tested for antibodies to SARS-CoV-2 to estimate the proportion of people who were previously infected. Statewide seroprevalence estimates for six U.S. states were compared with the computationally estimated cumulative incidence rates on the date one-week prior to the mid-points of the serum collection periods, accounting for time delays from infection to antibody detection.

Overall the described pipeline yielded accurate estimates of cumulative incidence when compared to seroprevalence rates, with exception of Utah (Fig. 2). The estimated cumulative incidence in New York by April 17, was 9.6% (5.2%–15.7%), which was in line with a seroprevalence of 14.0% (NYS), although the estimate had a large uncertainty originating in the wide confidence interval of the IFR estimate. The cumulative incidence rates in Washington state were 0.6% (0.5%–0.7%) and 1.9% (1.2%–2.9%) by March 21 and April 27, respectively, which were close to the seroprevalence of 1.1% and 2.1% surveyed in western Washington state measured one week later. The estimated incidence rate of 3.7% (1.9%–6.3%) in Connecticut by April 23, was in line with the seroprevalence of 4.9% in the first round, while the incidence estimate of 12.3% (6.1%–20.8%) by May 17, differed from seroprevalence 5.2% in the second round. As some studies reported that the antibodies decreased over time in some patients (14, 15), the second round seroprevalence rates in the four states seemed to be unstable. Indeed, the rates became even smaller than the first round in Utah and remained almost the same in Connecticut and Missouri. In comparison to the seroprevalence rates in the first round in Louisiana and Missouri, the cumulative incidence rates seemed to be slightly underestimated. In Utah, the estimates and seroprevalence rates showed prominent discrepancy, where the significantly low incidence estimate by April 20 suggested a possibility of under-reported death tolls.

**Figure 2.**
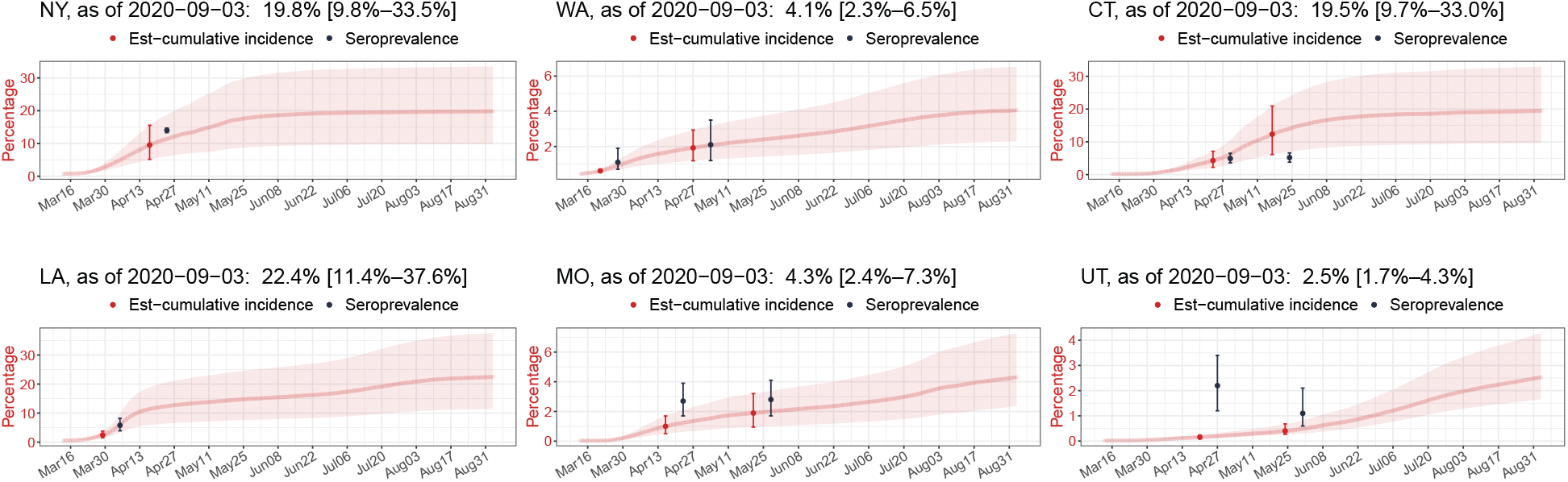
Validation of prediction framework using seroprevalence rates in six U.S. states. Seroprevalence rates (black) are overlaid on computationally estimated time courses of cumulative incidence rates (red) from March 13 to September 3, 2020, for New York, Washington state, Connecticut, Louisiana, Missouri, and Utah from upper-left to lower-right. The indicated date of the seroprevalence rate is the mid-point of the serum collection period. The corresponding cumulative incidence estimate is on the date one-week prior to the date of the seroprevalence rate to account for time delays from infection to antibody detection. Error bars and shaded bands indicate 95% confidence intervals.

Applied across countries and U.S. states, the proposed framework estimated actual time courses of new infections and currently infected cases. In early April, the U.S. reported ∼30,000 daily confirmed cases. In striking contrast, the proposed estimation suggested a number of actual daily cases of more than 400,000, showing that the daily ascertainment at that time was less than 10% (Fig. 3A). As of September 3, 2020, 0.9% (0.5%–1.6%) of the U.S. population was estimated to be currently infected. In Brazil, the under-reporting was also severe early in the pandemic, but gradually improved as in the U.S. As a result, the peak in actual daily cases seemed to have occurred between June 1 and June 8, 2020, reaching nearly 250,000 cases in contrast to ∼25,000 confirmed daily cases (Fig. 3B). This time of the peak in new infections was earlier than the peak in confirmed cases, which fell between July 27 to August 3. The currently infected cases in Brazil were estimated to be 2.3% (1.1%–3.9%) of the total population as of September 3, 2020. Among U.S. states, Louisiana showed the highest estimated fraction of currently infected people, 6.9% (3.7%–11.3%) as of September 3, 2020 (Fig. 3C). The first peak in the daily new cases in Louisiana was allegedly ∼1,500 around April 6, but the actual new cases at that time were estimated to be more than 30,000, indicating the severity of under-reporting in Louisiana during the month of April.

**Figure 3.**
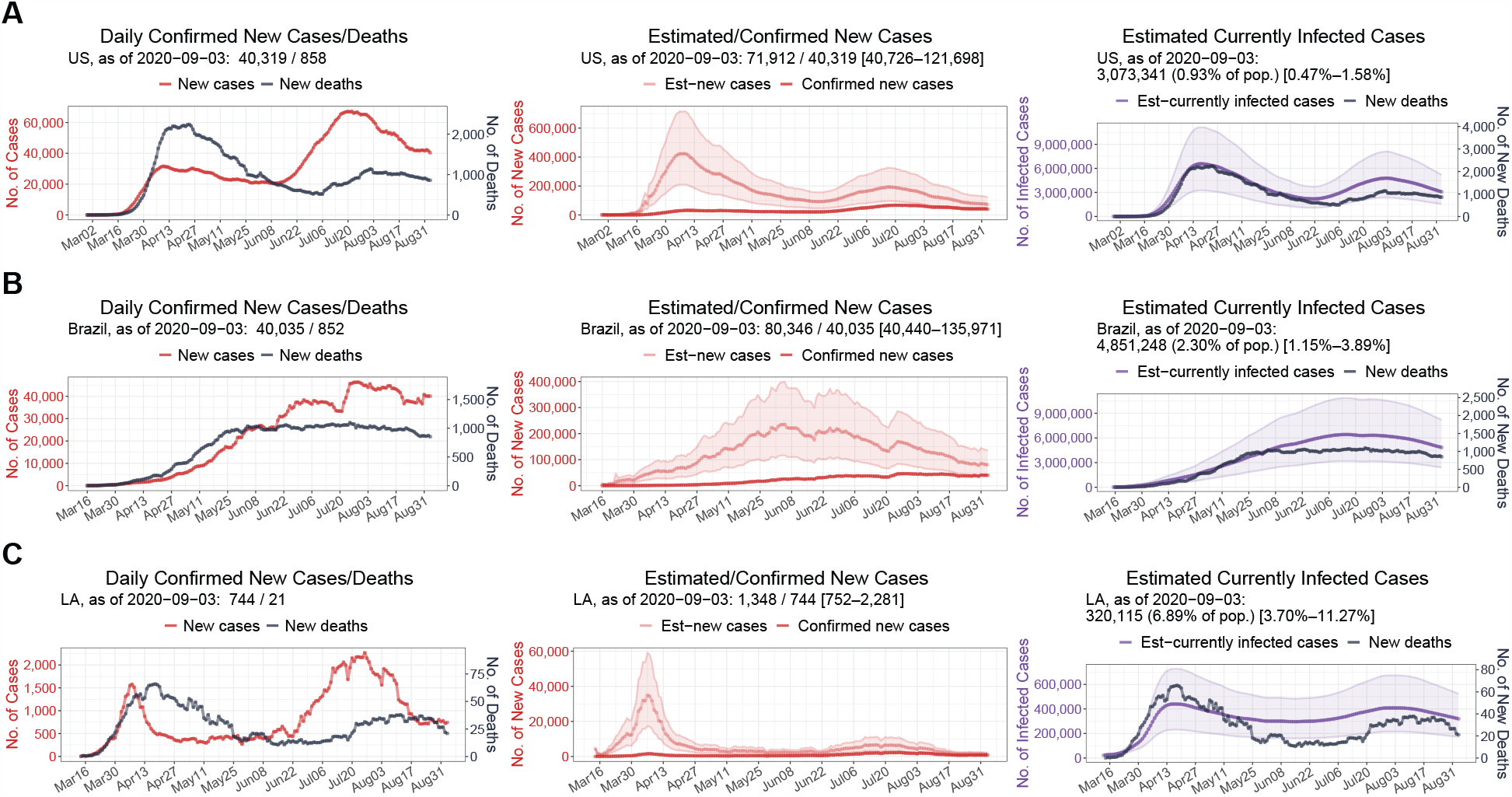
Estimated time courses of actual new cases and current infections. 7-day rolling-averaged counts of daily confirmed new cases and deaths (left) until September 3, 2020, for the U.S. (**A**), Brazil (**B**), and Louisiana (**C**). An estimate of new cases (middle) is the under-reporting-adjusted number of newly infected individuals each day. An estimate of current infections (right) is the under-reporting-adjusted number of infected individuals who have not yet been recovered or deceased. Shaded bands indicate 95%-confidence intervals.

The severe under-ascertainment was universal across the 50 countries with the most confirmed cases and 50 U.S. states. The ascertainment rates for the whole period until September 3, 2020, widely varied from 5% in Italy to 99% in Qatar, and from 8% in Connecticut to 71% in Alaska (Fig. 4A). Among them, 25 countries, 19 U.S. states, and Washington D.C. showed an ascertainment rate less than 20% for the entire time of the pandemic. Focusing on only the past two weeks the ascertainment rates unfortunately have not improved much in these countries, while the recent rates of U.S. states increased overall as of September 3, 2020. Interestingly, many of the countries with high ascertainment rates from the beginning of the outbreak were the ones that previously experienced Middle East respiratory syndrome coronavirus.

**Figure 4.**
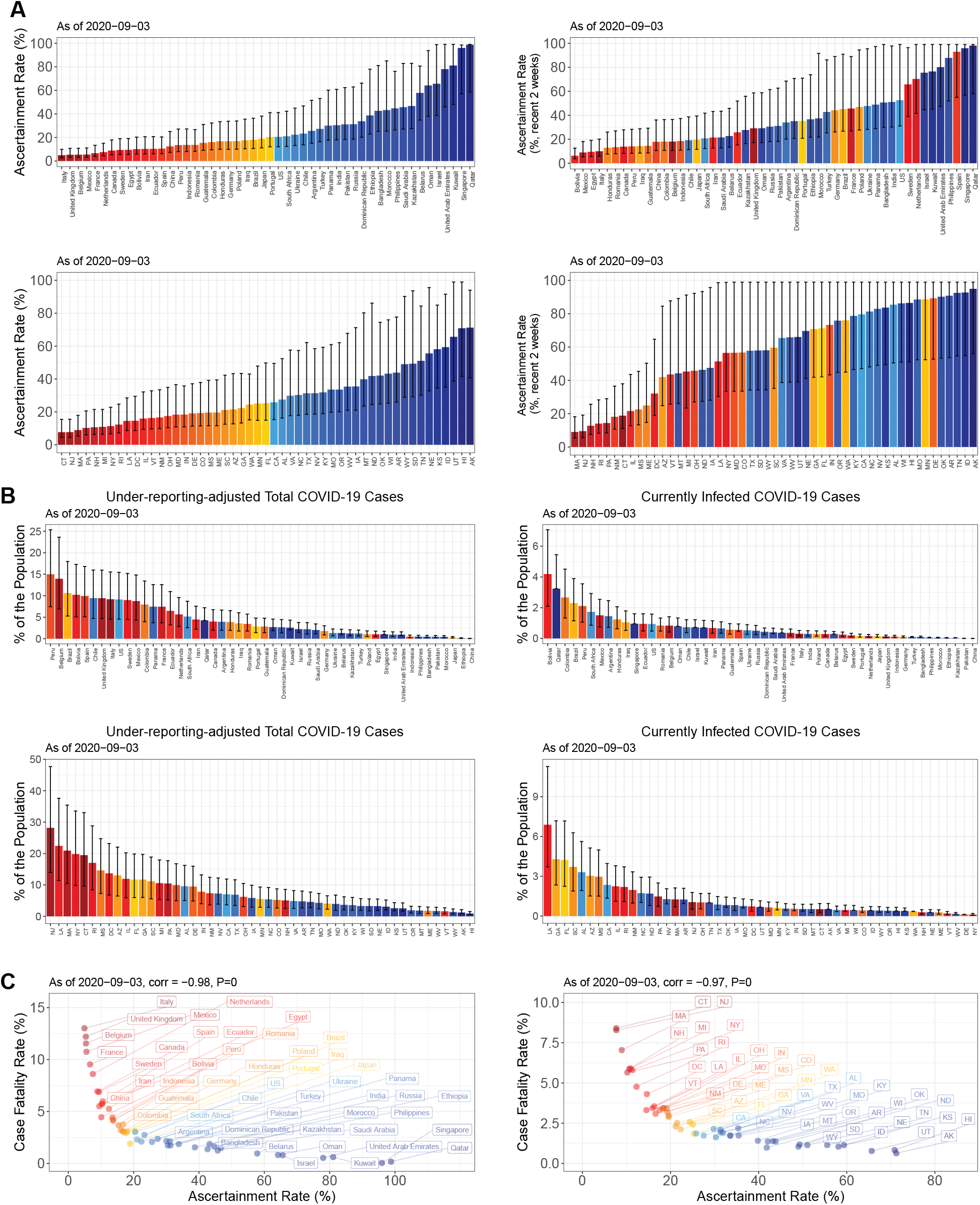
Estimates of ascertainment rates, cumulative incidence rates, and actual fractions of current infections in 50 countries and 50 U.S. states. (**A**) Estimates of ascertainment rates for the whole period until September 3, 2020 (left), and recent ascertainment rates (August 21–September 3, 2020) (right), in 50 countries with the most confirmed cases (upper) and 50 U.S. states (lower). (**B**) Cumulative incidence rates (left), and percentages of currently infected individuals in each population (right) in the 50 countries (upper) and 50 U.S. states (lower). Error bars indicate 95%-confidence intervals. (**C**) Scatter plots between the crude case-fatality-rates and the ascertainment rates for the 50 countries (left) and 50 U.S. states (right). Spearman rank correlations and their P-values are shown.

The under-reporting adjustment allowed us to monitor the actual severity of the virus spread across countries and U.S. states, and especially the estimated sizes of currently infected populations helped to identify fast-changing COVID-19 hotspots. In Peru, Belgium, and Brazil, more than 10% of the population were estimated to be once infected as of September 3, 2020 (Fig. 4B). Across U.S. states, the cumulative incidence rates ranged from 28.2% (14.0%–47.7%) in New Jersey to 0.9% (0.6%–1.5%) in Hawaii (Fig. 4B). As of September 3, 2020, COVID-19 hotspots among U.S. states were estimated to be Louisiana, Georgia, and Florida, where currently infected cases were estimated to be more than 4%.

The estimated fractions of current infections differentiated New Jersey from New York, both of which experienced severe early outbreaks (Fig. 4B). The confirmed new cases per 100,000 population were 3.8 in New Jersey and 3.7 in New York as of September 3, 2020, suggesting that the virus spread was under control in both states. However, because of the differences in recent ascertainment rates between the two states (Fig. 4A) the fractions of currently infected people were 1.05% (0.52%–1.78%) in New Jersey and 0.12% (0.06%–0.20%) in New York, as of September 3, 2020. This reveals New Jersey as still with a considerable infected population whereas New York has become one of the safest states.

Since the beginning of the COVID-19 pandemic, the Case-Fatality-Rates (CFRs) have displayed huge differences between countries, adding confusion to how deadly SARS-CoV-2 is. The crude CFRs, which are the ratios of total confirmed deaths to total confirmed cases, ranged from 13.0% in Italy to 0.05% in Singapore as of September 3, 2020. Our analysis now reveals that the variation on CFR reports across the countries and U.S. states is primarily associated with the massive differences in ascertainment rates between the locations (Fig. 4C). The Spearman rank correlations between CFR and ascertainment rate were −98% and −97% (P-values < 0.0001) for the analyzed countries and the U.S. states, respectively. After adjustment for the under-reporting, the inferred IFRs, which were based on the assumed IFR 0.66%, did not correlate with the ascertainment rates (Fig. S2). Thus, a high CFR in a region is shown to be a result of severe under-reporting of the cases.

This study demonstrates that severe under-ascertainment has obscured the true severity of widespread COVID-19 all over the world. In the majority of the 50 countries, actual cumulative cases were estimated to be 5–20 times greater than the confirmed cases. Given that the confirmed cases only capture the tip of the iceberg in the middle of the pandemic, the estimated sizes of current infections in this study provide crucial information to determine the regional severity of COVID-19 that can be misguided by the confirmed cases.

It is a challenging task to estimate actual numbers of COVID-19 infections based on under-reported limited data, especially to make a framework applicable for many regions that have displayed diverse dynamic patterns in the infections and ascertainment rates. As the pandemic progresses, the pipeline would need to be adapted to the increasing complexity of the infection data. The proposed estimation heavily relies on the published estimate of the IFR that has large estimation uncertainty. The estimates of actual cases would become more accurate if the IFR estimate is optimized to a specific region and its uncertainty can be reduced. The pipeline only takes the simple input of the confirmed cases and deaths. Depending on available datasets in each region, the estimation of actual cases can be improved by augmenting more information such as daily positivity rates of diagnostic testing or daily hospitalized cases.

## Data Availability

All code, daily updated estimates, and their visualizations are freely available at a GitHub repository (https://github.com/JungsikNoh/COVID19_Estimated-Size-of-Infectious-Population).

## Author contributions

J.N. conceived of the study, developed the method, and performed the analysis. J.N. and G.D. wrote the manuscript.

## Supplementary Materials

### Materials and Methods

#### 1. Initialization of actual time courses of new cases and current infections

The proposed framework provides estimates of time courses of actual new cases and current infections based on daily reported counts of confirmed positive tests and deaths in a particular region and published estimates of key pandemic parameters such as the infection-fatality-rate (IFR) and the mean duration from infection to death and recovery. For the purpose of this study data of daily confirmed cases and deaths for countries and U.S. states were taken from the repository by the Center for Systems Science and Engineering (CSSE) at Johns Hopkins University (1), and the COVID Tracking Project (2), respectively. Upon availability of other more granular data sets, the proposed framework can also be applied to smaller populations. The data of the country population as of July 2019 was from the United Nations (3), and the population data for U.S. states as July 2019 was from the U.S. Census Bureau (4).

To begin with, the counts of daily confirmed cases and deaths were averaged in a 7-day rolling window to remove weekend effects, which tend to yield systematic drops in the Saturday and Sunday values. To obtain initial guesses on actual counts of daily new cases and recoveries, we first combined the daily death counts and the IFR estimate (*IFR*_0_ = 0.66%) that was presented by Verity et al. (5) as follows: Let *AC*_*t*_, *CC*_*t*_, *I*_*t*_, *D*_*t*_, *R*_*t*_ denote the number of actual new cases, confirmed new cases, currently infected cases, new deaths, and new recoveries on day t, respectively. Using the mean duration estimates (5), we assumed that one new death on day t implied 1/*IFR*_0_ new actual cases on day (*t* − *d*_1_), *d*_1_ = 18, and 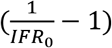 new recoveries on day (*t* + *d*_2_), *d*_2_ = 7. From this follows

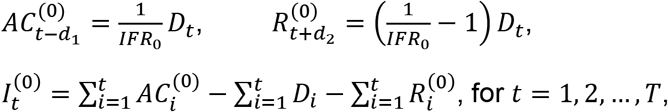

where 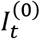 is the derived initial estimate of the currently infected cases on day t. These initial time courses led to two other initial estimates for daily ascertainment rates (*A*_*t*_), and daily ratios of the confirmed new cases to currently infected cases, referred to as detected transmission rates (*DT*_*t*_):

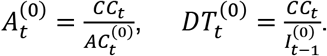

Both ratio estimates displayed a common increasing trend in many countries, probably indicating that the under-ascertainment was gradually improving as the testing capacities were increasing. The common trend could be exploited to obtain better estimates of the daily ascertainment rates.

#### 2. Expectation-maximization iterations to update latent time courses

An expectation-maximization (EM) algorithm was implemented to update the latent time courses involved in actual infections. To first extract the temporal trends of the estimated daily ascertainment rates and detected transmission rates, the two rate time courses were spline-smoothed. Since the noise-levels around the temporal trends were different depending on regional population sizes, infection dynamics, and test reporting schemes, we applied multiple levels of smoothness and selected the optimal level after the complete EM computation as discussed below. For smoothing, we applied the R (6) function *smooth*.*spline()*. Since the smoothed estimates of the detected transmission rates and ascertainment rates (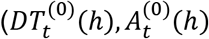, *h* is a smoothness parameter) possessed a common trend, the estimated ascertainment rates could be improved by augmenting the information from 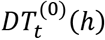. The following regression model was applied to find a functional relation from 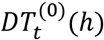 to 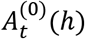:

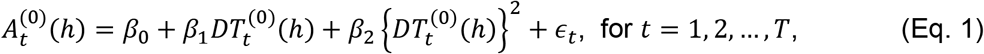

where the regression coefficients were estimated with constraints of *β*_1_, *β*_2_ ≥ 0 using an R function *penalized()*. Then, the estimated coefficients 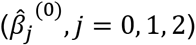 were used to update the initial ascertainment rates:

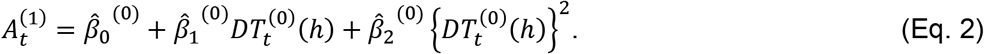

The above EM steps enabled iterative updates of the latent time courses on actual infections. For each smoothness parameter *h*, the following EM algorithm was applied to obtain converged estimates (Fig. S1A).

##### Algorithm 1. EM iterations to update the latent time courses

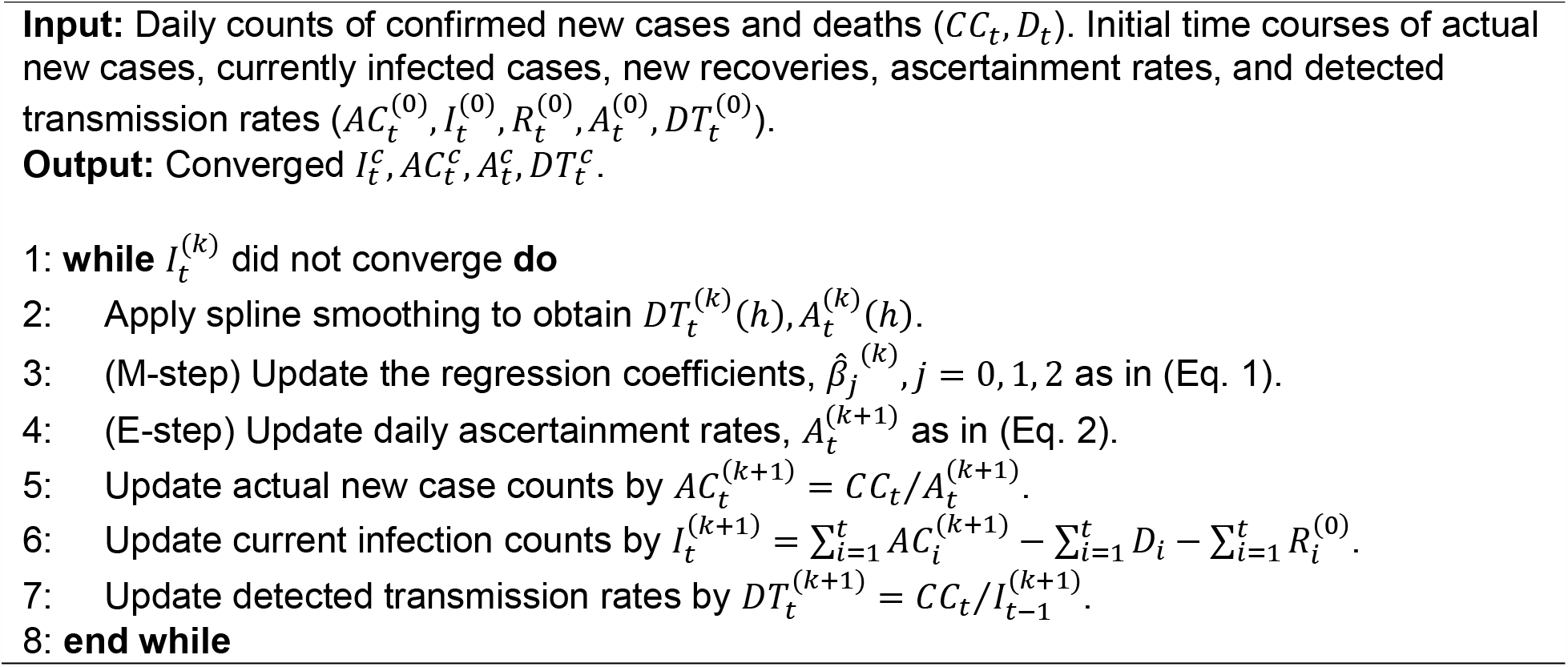

### 3. Optimization toward the smallest variation of daily death rates

After completing EM iterations with multiple smoothness parameters (*h*), the converged time courses of currently infected cases were assessed by using corresponding daily rates of deaths among the infected cases:

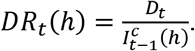

The framework selected the smoothness parameter value (*ĥ*) and the final estimate of current infections 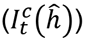 that produced the smallest coefficient of variation (CV) of *DR*_*t*_(*h*) over time. Underlying this choice is the assumption that increasing variation in daily death rates would be less plausible.

To obtain 95%-confidence intervals (CIs) of the current infection estimates, the same initialization and EM iterations were applied using the lower/upper limits of the IFR estimate and the selected smoothness (*ĥ*). The workflow of the initialization, EM iterations, and calculating CIs are illustrated with the case of the U.S. time courses (Fig. S1B).

### 4. Data and code availability

The estimates of actual new infections and currently infected cases have been updated daily for 50 countries with the most confirmed cases and 50 U.S. states since August 12, 2020, in the GitHub repository (https://github.com/JungsikNoh/COVID19_Estimated-Size-of-Infectious-Population). All code, daily updated estimates, and visualizations are publicly available in this online repository.

## Figure Legends

**Fig. S1.**
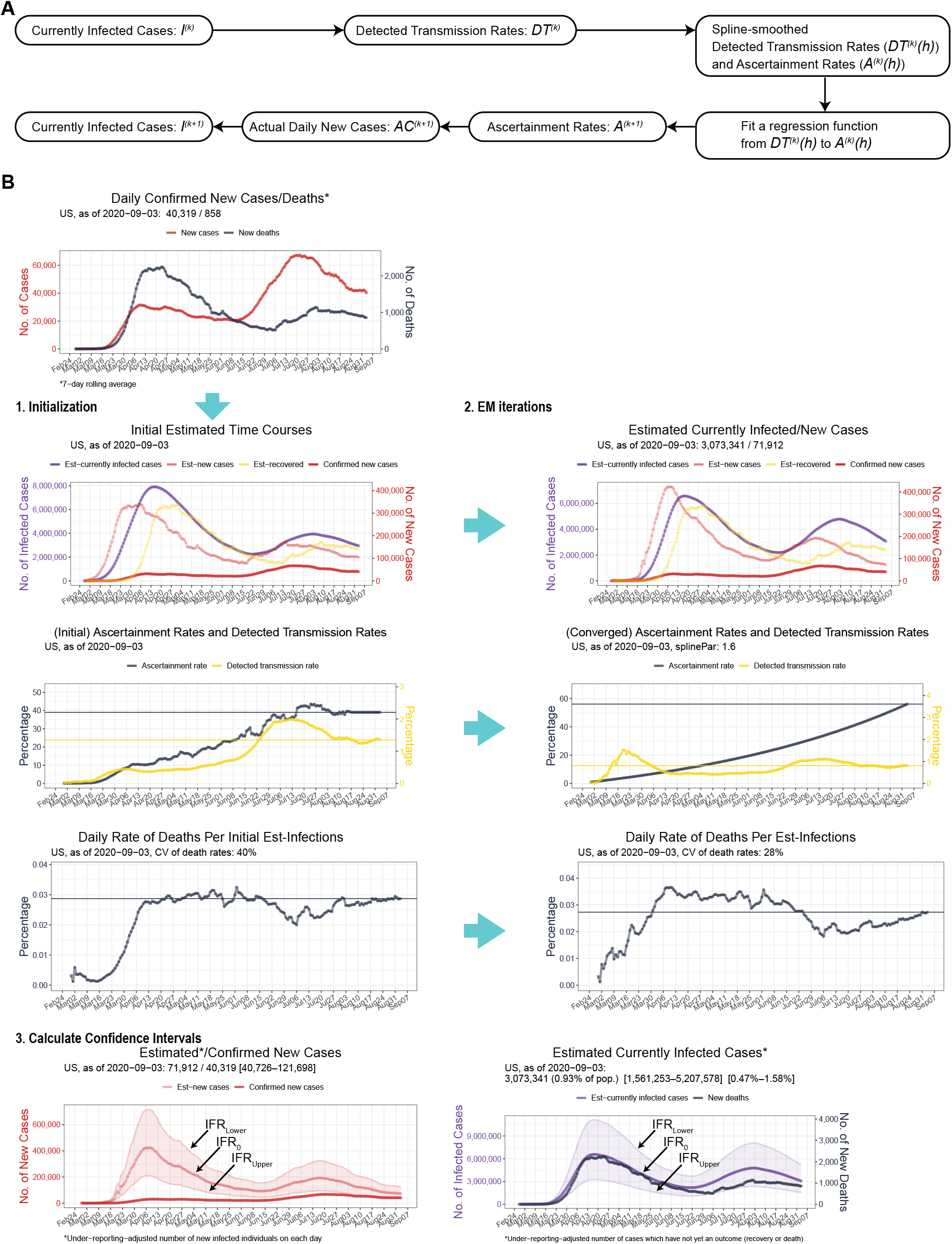
Workflow to estimate time courses of actual infections. (**A**) Expectation-maximization (EM) iteration to update latent time courses involved in actual infections. (**B**) Workflow of initialization, EM iterations, and calculation of confidence intervals.

**Fig. S2.**
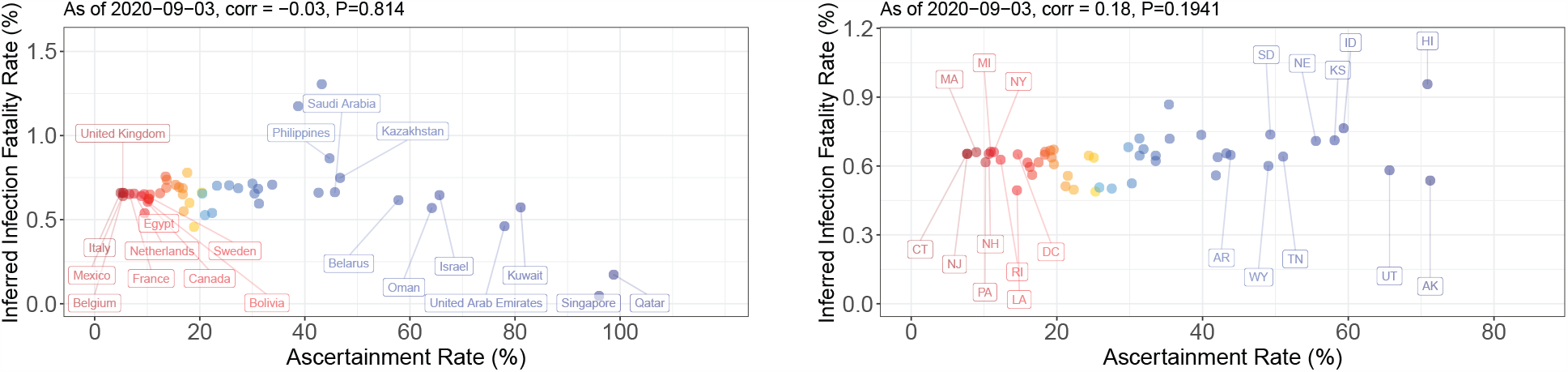
Inferred infection-fatality rates and ascertainment rates. Scatter plots between the inferred infection-fatality-rates (IFR) and the whole period ascertainment rates for the 50 countries (left) and 50 U.S. states (right). The inferred IFR is the ratio of total confirmed deaths to the under-reporting-adjusted total number of cases on a date 18-day before, accounting for the mean duration from infection to death. Spearman rank correlations and their P-values are shown.

